# Emergence of an early SARS-CoV-2 epidemic in the United States

**DOI:** 10.1101/2021.02.05.21251235

**Authors:** Mark Zeller, Karthik Gangavarapu, Catelyn Anderson, Allison R. Smither, John A. Vanchiere, Rebecca Rose, Daniel J. Snyder, Gytis Dudas, Alexander Watts, Nathaniel L. Matteson, Refugio Robles-Sikisaka, Maximilian Marshall, Amy K. Feehan, Gilberto Sabino-Santos, Antoinette R. Bell-Kareem, Laura D. Hughes, Manar Alkuzweny, Patricia Snarski, Julia Garcia-Diaz, Rona S. Scott, Lilia I. Melnik, Raphaëlle Klitting, Michelle McGraw, Pedro Belda-Ferre, Peter DeHoff, Shashank Sathe, Clarisse Marotz, Nathan Grubaugh, David J. Nolan, Arnaud C. Drouin, Kaylynn J. Genemaras, Karissa Chao, Sarah Topol, Emily Spencer, Laura Nicholson, Stefan Aigner, Gene W. Yeo, Lauge Farnaes, Charlotte A. Hobbs, Louise C. Laurent, Rob Knight, Emma B. Hodcroft, Kamran Khan, Dahlene N. Fusco, Vaughn S. Cooper, Phillipe Lemey, Lauren Gardner, Susanna L. Lamers, Jeremy P. Kamil, Robert F. Garry, Marc A. Suchard, Kristian G. Andersen

## Abstract

The emergence of the early COVID-19 epidemic in the United States (U.S.) went largely undetected, due to a lack of adequate testing and mitigation efforts. The city of New Orleans, Louisiana experienced one of the earliest and fastest accelerating outbreaks, coinciding with the annual Mardi Gras festival, which went ahead without precautions. To gain insight into the emergence of SARS-CoV-2 in the U.S. and how large, crowded events may have accelerated early transmission, we sequenced SARS-CoV-2 genomes during the first wave of the COVID-19 epidemic in Louisiana. We show that SARS-CoV-2 in Louisiana initially had limited sequence diversity compared to other U.S. states, and that one successful introduction of SARS-CoV-2 led to almost all of the early SARS-CoV-2 transmission in Louisiana. By analyzing mobility and genomic data, we show that SARS-CoV-2 was already present in New Orleans before Mardi Gras and that the festival dramatically accelerated transmission, eventually leading to secondary localized COVID-19 epidemics throughout the Southern U.S.. Our study provides an understanding of how superspreading during large-scale events played a key role during the early outbreak in the U.S. and can greatly accelerate COVID-19 epidemics on a local and regional scale.

## Introduction

In December 2019, SARS-CoV-2 was first identified in pneumonia cases in Wuhan, China^1,2^. Initially, community transmission was confined to China, but in late February 2020 large-scale outbreaks were increasingly detected in Europe and the Middle East^3,4^. Although SARS-CoV-2 was initially detected in the United States (U.S.) in January 2020^5^, the majority of early COVID-19 cases were associated with travel from high-risk countries or close contact with travelers^6^.

By late February, wide-spread community transmission of SARS-CoV-2 in the U.S. was identified in Washington state^7^, New York City^8^, and Santa Clara County in California^9^, but it is estimated that local transmission in the U.S. started earlier and was more wide-spread than recognized at the time^10,11^. Elsewhere, outside of these early virus ‘hotspots’ in the U.S., transmission of SARS-CoV-2 occurred mostly silently due to lack of testing until the second week of March^10,12,13^. It seems likely that large-scale events in this period dramatically accelerated early SARS-CoV-2 transmission and that subsequent interstate seeding amplified the COVID-19 epidemic in the U.S..

More than one million people from all over the U.S. were drawn to the Mardi Gras parades in New Orleans starting on February 14^th^ and culminating on February 25^th^, 2020 (Mardi Gras day / “Fat Tuesday”). The timing and the scale of this event, as well as the absence of any meaningful mitigation efforts (in agreement with official guidelines), provides a unique opportunity to investigate how large-scale events can accelerate SARS-CoV-2 transmission and amplify local outbreaks during the ongoing pandemic. To investigate this, we sequenced SARS-CoV-2 from cases in New Orleans and other locations in Louisiana and compared them with SARS-CoV-2 genomes from the U.S. and globally to reconstruct the timing, origin, and emergence of the virus in Louisiana. By integrating genomic, epidemiological, and mobility data we show that SARS-CoV-2 overdispersion during Mardi Gras greatly accelerated the early outbreak in New Orleans and seeded the virus to other parts of Louisiana and nearby states. Our findings suggest that large-scale events in the beginning of 2020 may have contributed significantly to SARS-CoV-2 transmission early in the COVID-19 epidemic in the U.S. and that without widespread availability of vaccination and testing, large gatherings of people without strict control efforts will continue to amplify the COVID-19 pandemic.

## Results

### SARS-CoV-2 was likely introduced into Louisiana via domestic travel

To understand the early emergence of SARS-CoV-2 in Louisiana, we investigated epidemiological, genomic, and travel data of SARS-CoV-2 during the first wave of the epidemic (March 9^th^ - May 15^th^). We found that SARS-CoV-2 in Louisiana displayed little genetic diversity compared to other states and was likely introduced from a domestic source.

Using aggregated parish-level COVID-19 case data^14^, we analyzed reported cases and deaths during the first wave of the epidemic in Louisiana. The first reported case of COVID-19 in Louisiana was detected on March 9^th^ 2020 and the epidemic rapidly increased with reported cases reaching a peak on April 4^th^ (**Figure 1A**). While COVID-19 cases were reported throughout Louisiana during the first wave, the New Orleans-Metairie metropolitan statistical area (MSA; henceforth referred to as New Orleans) accounted for more than 54.9% of all deaths in the period up until May 1^st^ (**Figure S1**) and was the focal point of the epidemic in Louisiana.

**Figure 1.**
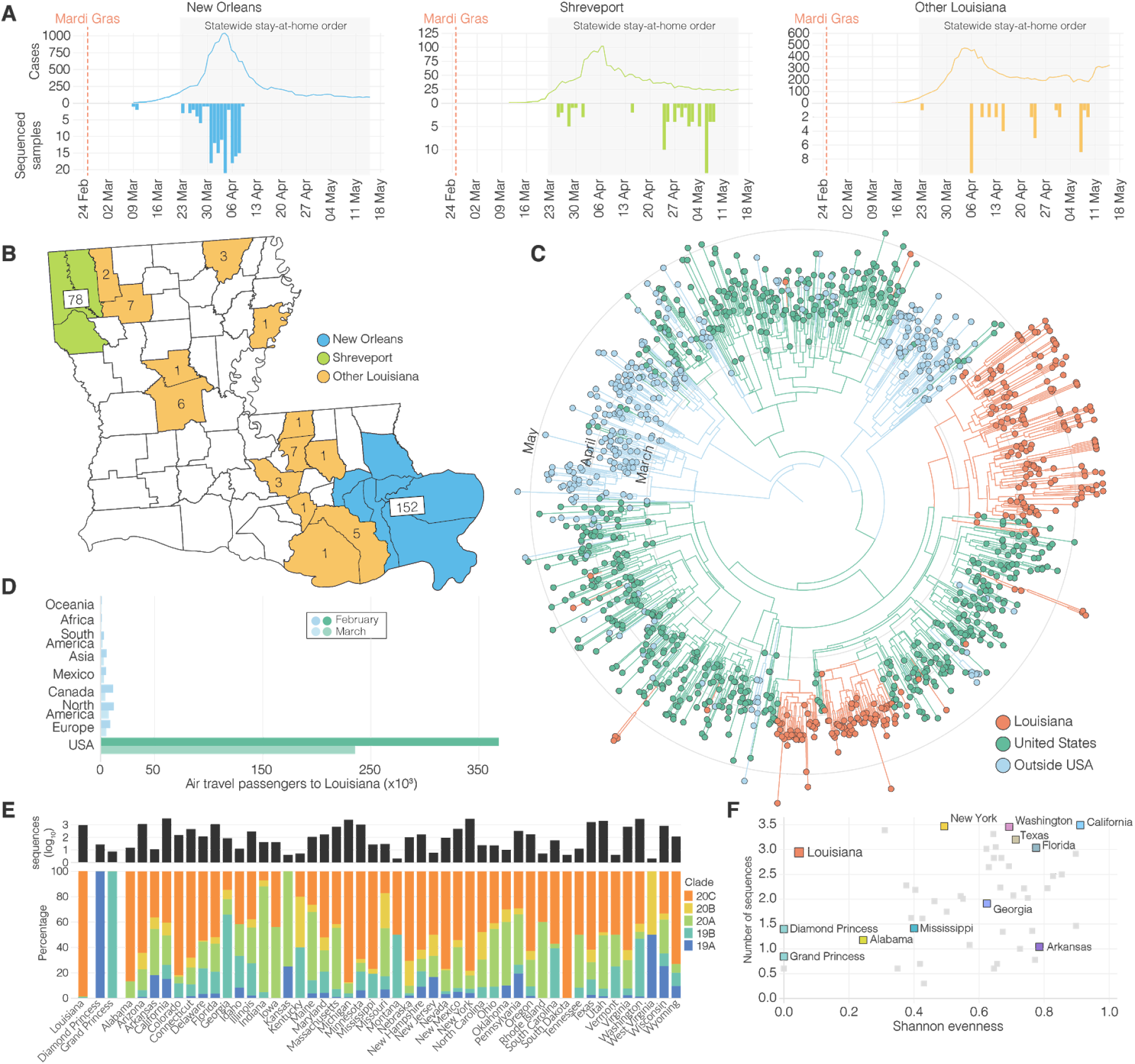
SARS-CoV-2 epidemiology in Louisiana. (**A**) Epidemiological curve and number of sequenced samples in New Orleans, Shreveport and other parishes in Louisiana. (**B**) Sampling location of sequenced SARS-CoV-2 samples in Louisiana: New Orleans metro area (blue), Shreveport metro area (green), and other parishes in Louisiana (orange). (**C**) Maximum clade credibility tree of SARS-CoV-2 sequences sampled from Louisiana, U.S. and outside the U.S.. (**D**) Domestic and international air travel passenger volumes to Louisiana in February and March. (**E**) Relative NextStrain clade prevalence per U.S. state up until May 15th (bottom). Number of sequences per U.S. state up until May 15th (top). (**F**) Shannon evenness of NextStrain clades per U.S. in relation to available sequences.

Early SARS-CoV-2 epidemics in New York and the West Coast were seeded by international introductions from Europe and Asia, respectively^7^. However, the source of many other local epidemics in the U.S., including the one in Louisiana, is unknown. To determine whether the emergence of SARS-CoV-2 in Louisiana originated from a domestic or international source, we sequenced 235 SARS-CoV-2 virus genomes collected from COVID-19 patients in New Orleans, Shreveport (Shreveport-Bossier City, LA MSA) and other parishes in Louisiana (**Figure 1A, B**). We reconstructed phylogenetic trees together with 1,263 whole genome sequences that were representative of the global SARS-CoV-2 sequence diversity between January and May, 2020. We found that the lineages responsible for the first wave in Louisiana all closely resembled SARS-CoV-2 sequences sampled within the U.S., suggesting that the epidemic in Louisiana was seeded from a domestic source (**Figure 1C**).

To further investigate the origin of the SARS-CoV-2 introduction into Louisiana, we investigated domestic and foreign air travel into Louisiana, and found that in February, 360,000 passengers arrived from within the U.S., while only 40,000 international travelers were reported (**Figure 1D**). In particular, we found that travel from Europe and Asia, where the majority of SARS-CoV-2 transmissions occurred in February, accounted for less than 5% of all travel movements to Louisiana (**Figure 1D**). Consistent with our phylogenetic analysis, the travel data strongly suggest that the COVID-19 epidemic in Louisiana was due to seeding from domestic sources of SARS-CoV-2, and, unlike New York^8^ and Washington^7^, not the result of importations from Europe, Asia or other foreign countries.

### Early SARS-CoV-2 transmission in Louisiana predominantly originated from a single introduction

Unrestricted domestic travel in the U.S. in February, 2020 and associated large travel volumes likely facilitated the emergence of SARS-CoV-2 in Louisiana. To investigate how many times SARS-CoV-2 was introduced into Louisiana, we first conducted a high-level genomic analysis by comparing NextStrain clade distributions of all available SARS-CoV-2 sequences from the continental U.S. up until May 15^th^, 2020. We found that SARS-CoV-2 sequences from Louisiana almost exclusively belonged to a single clade, 20C (**Figure 1E**). In other U.S. states with more than 10 sequences available, including neighboring states of Louisiana, we observed the co-circulation of multiple clades at more equal frequencies than in Louisiana (**Figure 1E, F**). In fact, we found that the genetic diversity of SARS-CoV-2 in Louisiana strongly resembled outbreaks on cruise ships (**Figure 1E, F**). These findings suggest that, like on the Diamond Princess and Grand Princess cruise ships^9,15^, SARS-CoV-2 in Louisiana most likely originated from a single source.

To further support these findings, we reconstructed a maximum likelihood tree of our SARS-CoV-2 genomes from Louisiana together with a representative selection of 1,399 clade 20C sequences collected across the U.S. (**Figure 2A**). We found that, within clade 20C, the majority of SARS-CoV-2 sequences in Louisiana belonged to a single cluster (“Louisiana clade”; **Figure 2A, B**), which is characterized by a single defining nucleotide mutation (C27964T; **Figure 2A**). Within the Louisiana clade, we identified three additional subclades supported by single nucleotide mutations, but the Louisiana clade was otherwise strongly dominated by polytomies, consistent with rapid local transmission (**Figure 2A**). Outside the main Louisiana clade, we found ten singleton sequences, but these either resulted in very limited or no onward transmission and likely did not contribute substantially to the overall SARS-CoV-2 transmission during the first wave (**Figure 2A**). The clustering of SARS-CoV-2 sequences within a single well-supported Louisiana clade strongly suggests that a single introduction was responsible for the vast majority of transmission events during the first wave of the epidemic in Louisiana.

**Figure 2.**
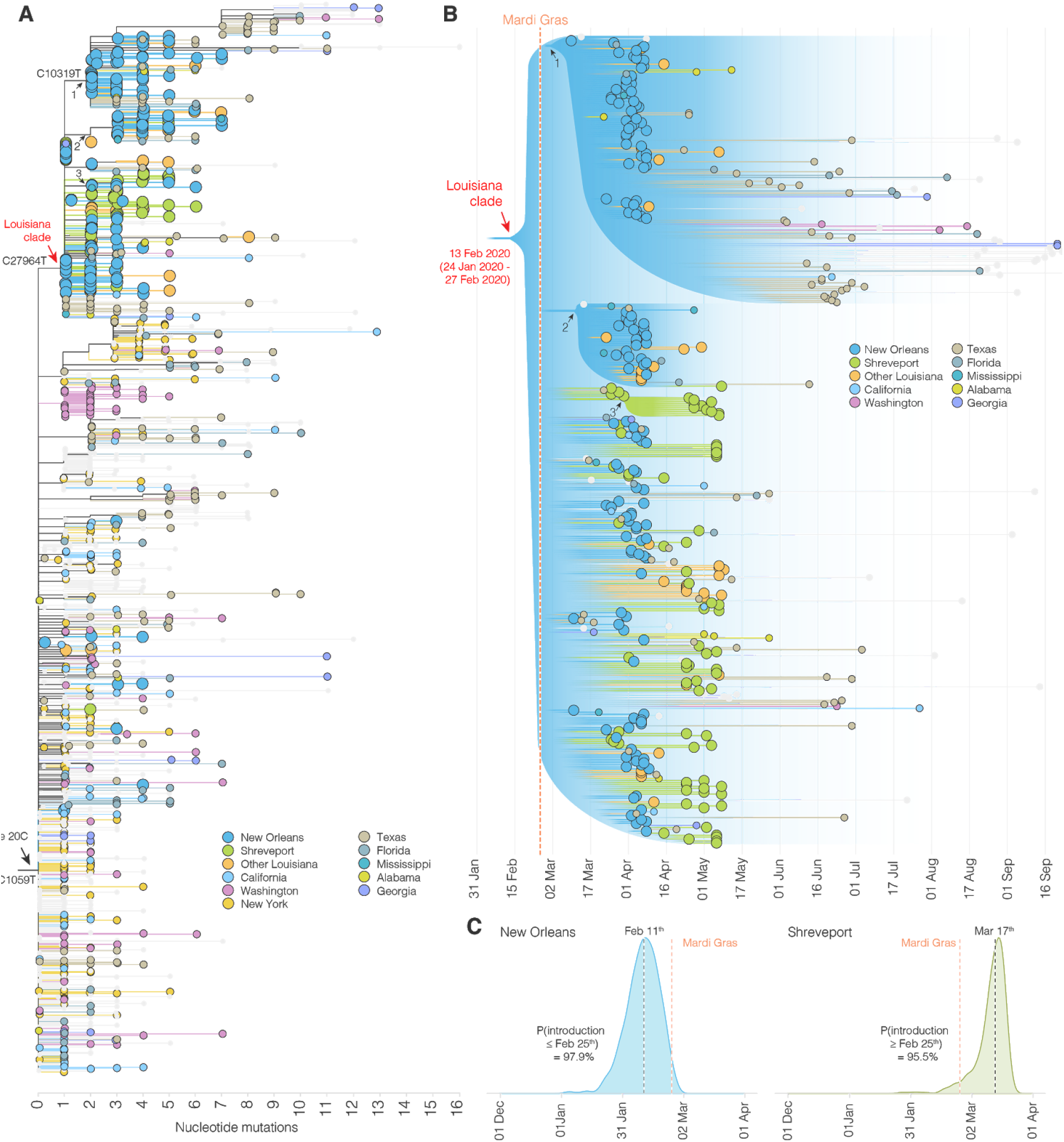
Phylogenetic analysis of SARS-CoV-2 in Louisiana. (**A**) Maximum likelihood tree of SARS-CoV-2 genomes sequenced in Louisiana and other parts of the U.S.. U.S. states that are not color-coded are indicated in grey. Arrows indicate clades. (**B**) Illustration of maximum clade credibility tree. Gradients are used to illustrate uncertainty in the topology and node heights. Numbered arrows are nodes with a relatively high posterior support and correspond to the arrows in panel A. The red colored arrow indicates the most recent common ancestor of SARS-CoV-2 in Louisiana. (**C**) Posterior distribution of the first emergence into New Orleans (blue) and Shreveport (green). The time of the first state change to New Orleans and Shreveport along the phylogenetic tree of each posterior sample was computed and the posterior distribution was learned by summarizing across all the posterior samples.

### SARS-CoV-2 likely emerged in Louisiana prior to the Mardi Gras festival

Both the timing and the onset of the COVID-19 epidemic in New Orleans as well as media reports (**Table S1**) suggest that Mardi Gras, which culminated in large parades on Mardi Gras day on February 25^th^, 2020, may have played a role in the spread or emergence of SARS-CoV-2 in Louisiana. It is unclear, however, if SARS-CoV-2 was introduced during Mardi Gras or if local transmission was already ongoing prior to the festival. To evaluate when SARS-CoV-2 started circulating in Louisiana, we created time-aware phylogenies to estimate the median time to the most recent common ancestor (TMRCA) for the Louisiana clade, which indicates the likely start of sustained local transmission^16,17^. We found that the posterior median TMRCA of the Louisiana clade was February 13^th^ (95% highest posterior density [HPD] interval: January 24^th^ 2020 - February 27^th^ 2020), suggesting that low levels of local SARS-CoV-2 transmission within Louisiana was likely already ongoing prior to Mardi Gras (**Figure 2B**).

To further investigate potential local transmission prior to Mardi Gras, we determined the emergence of SARS-CoV-2 in Louisiana by inferring the timing of the first introduction (geographic state change, often called a Markov jump^18^) into New Orleans and Shreveport across our full model posterior distribution that includes uncertainty on the tree and model parameters. We estimated that SARS-CoV-2 lineages belonging to the Louisiana clade emerged in New Orleans with a median time of February 11^th^, 2020, which is two weeks before Mardi Gras day (Pr[introduction < February 25^th^] = 97.9%), and, in confirmation, two days before our TMRCA estimates of sustained local transmission on February 13^th^ (**Figure 2B, C**). In Shreveport, we found that SARS-CoV-2 emerged noticeably later than in New Orleans, after Mardi Gras on March 17^th^ (Pr[introduction > February 25^th^] = 95.5%; **Figure 2C**). Combined, our phylodynamic analyses suggest that SARS-CoV-2 emerged and spread locally in New Orleans prior to Mardi Gras.

### Increased transmission rates in New Orleans suggest superspreading during Mardi Gras

Although we found that SARS-CoV-2 likely began spreading in New Orleans mid-February, 2020, the first official COVID-19 case was not reported until March 9^th^. This suggests that SARS-CoV-2 was likely spreading undetected and unmitigated during the large-scale gathering of people during Mardi Gras. To determine whether the festival may have accelerated the early COVID-19 epidemic in Louisiana, we modeled the number of likely daily cases using reported deaths (**Figure 3A**) and compared these with simulated case numbers that, based on our TMRCA estimates (**Figure 2C**), assumed local transmission started on February 13^th^, 2020 (**Figure 3B**). We found that the observed number of infections was substantially higher than the expected number of infections, suggesting superspreading during Mardi Gras (**Figure 3C**).

**Figure 3.**
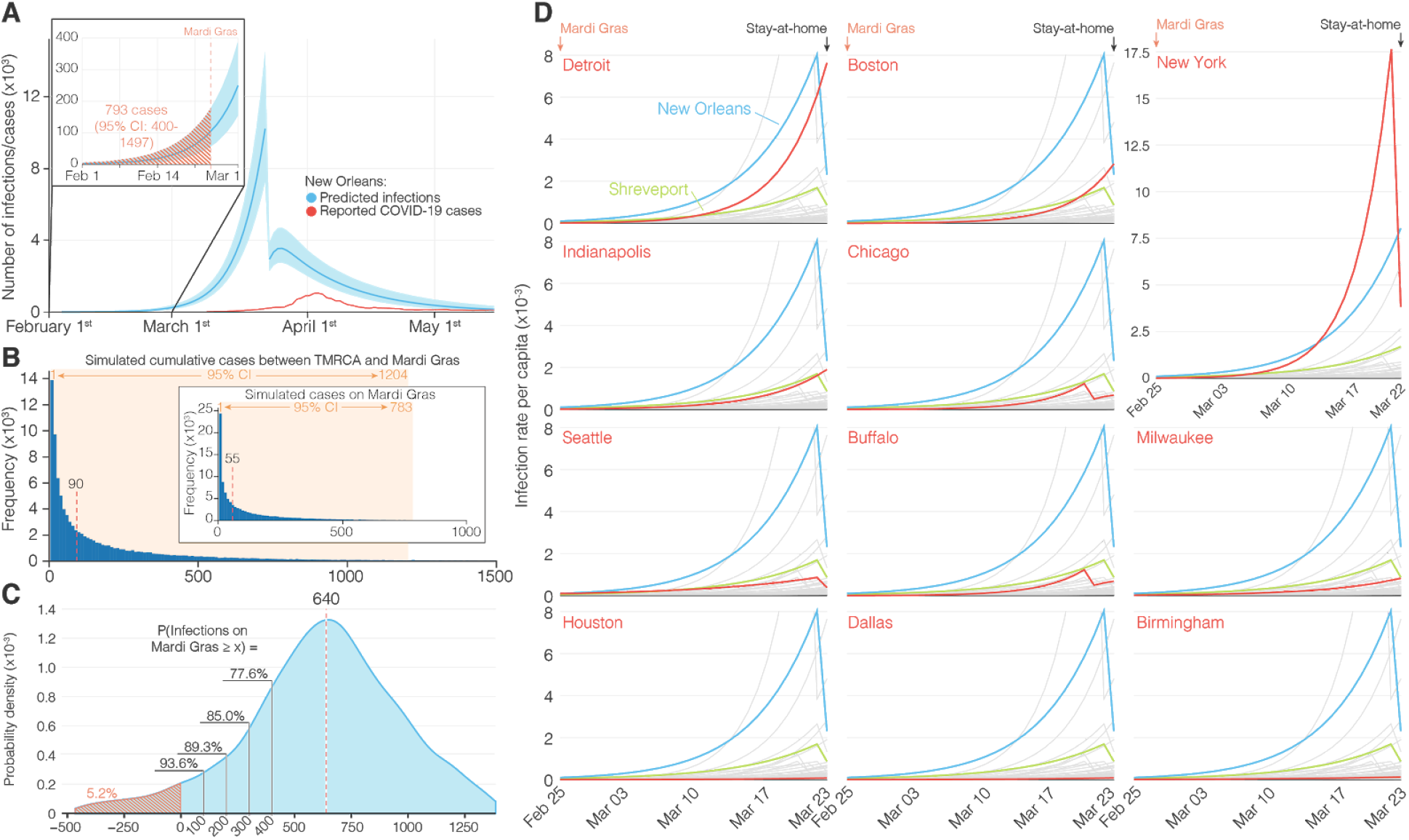
Acceleration of SARS-CoV-2 transmission during Mardi Gras. (**A**) Modeled incidence of SARS-CoV-2 in New Orleans based on registered COVID-19 deaths. The inset shows SARS-CoV-2 incidence in February and the hashed area indicates the cumulative number of COVID-19 cases up until Mardi Gras (February 25^th^, 2020). (**B**) Simulation of the cumulative number of infections between the TMRCA (February 13^th^) and the end of Mardi Gras. The inset shows a simulation of the number of infectious people on Mardi Gras day. The red dotted lines indicate the estimated median number of infections. (**C**) Probability density curve of the number of COVID-19 cases required on Mardi Gras day to recapitulate the epi curve in New Orleans. The red dotted line indicates the median number of cases. The hashed area is the probability that no increased transmission occurred during Mardi Gras. The black lines indicate the probability of accelerated transmission by 100, 200, 300, and 400 COVID-19 cases. (**D**) Modeled incidence of SARS-CoV-2 between Mardi Gras and the statewide stay at home order in Louisiana for New Orleans, Shreveport and 52 metro areas with a population of more than 1 million.

To estimate daily COVID-19 case numbers in the absence of reporting during February, 2020, we reconstructed the number of likely infections based on the number of reported deaths using a Bayesian regression model^19^. Since our model was not able to accommodate sudden increases in transmission that are typically associated with superspreading events^19^, we estimated the cumulative numbers up until Mardi Gras day on February 25^th^, 2020. We found that by Mardi Gras day, 793 (95% HPD: 400-1497) cumulative cases would have been required to align our model with the estimated daily number of SARS-CoV-2 infections during the first wave of the COVID-19 epidemic in New Orleans (**Figure 3A**). To estimate the likely number of infections in New Orleans between February 13^th^ (start of local transmission of the Louisiana clade; **Figure 2B**) and the end of Mardi Gras (February 25^th^), we simulated the number of cases using a negative binomial branching process model^20^. We estimated a total of 90 (95% confidence interval [CI]: 1-1204) cumulative infections occurred between February 13^th^ and Mardi Gras day (**Figure 3B**), which is substantially lower than the estimated 793 cumulative infections that would have been required to recapitulate the number of cases seen later in March (**Figure 3A**).

To estimate the number of likely SARS-CoV-2 infections during Mardi Gras, we calculated the median difference between our previously estimated cumulative infections up until Mardi Gras (793; **Figure 3A**) and the number of cases that were expected based on the start of local transmission on February 13^th^ (90; **Figure 3B**). We estimated that a median of 640 infections would have been required by Mardi Gras day to recapitulate our modeled epidemiological curve (**Figure 3C**), with only a 5.2 % probability that no transmission occurred at all during the festival. To better understand the magnitude of SARS-CoV-2 transmission during Mardi Gras, we randomly sampled the probability distribution of the modeled and simulated cases and calculated the probability of various transmission scenarios ranging from 100 to 400 additional infections during the festival. We found that at least 100 infections occurred during Mardi Gras with a 93.6% probability, and that at least 400 occurred with a 77.6% probability (**Figure 3C**). These findings suggest that superspreading very likely occurred during the festival resulting in hundreds of SARS-CoV-2 infections.

We hypothesized that superspreading during Mardi Gras should have resulted in a more rapid increase of early COVID-19 cases in New Orleans compared to other U.S. cities. To investigate this, we used a Bayesian regression model to estimate daily case numbers in New Orleans and other large population centers in the weeks after Mardi Gras until the statewide stay-at-home order in Louisiana on March 23^rd^, 2020^19^. We found that infection rates were substantially higher in New Orleans than in other large population centers, including cities with the next eight highest infection rates in the U.S (Detroit, Boston, New York, Indianapolis, Chicago, Seattle, Buffalo, and Milwaukee; **Figure 3D**). Since all of these population centers were located in the north and the west of the U.S., we also compared New Orleans with regional population centers in the South (Houston, Dallas, Birmingham, and Shreveport). We found substantially higher infection rates in New Orleans compared with these regional cities, indicating that infection rates in New Orleans were uniquely high in the Southern U.S. (**Figure 3D**). The increased rate of early COVID-19 cases in New Orleans suggests that superspreading occurred during Mardi Gras, which is in agreement with our previous analyses (**Figure 3A, B, C**).

### SARS-CoV-2 in Louisiana was highly similar to SARS-CoV-2 lineages circulating in Texas

Our analyses showed that SARS-CoV-2 was most likely introduced into Louisiana via domestic travel (**Figure 1C**). To more precisely determine the likely source of SARS-CoV-2 into Louisiana, we performed Bayesian phylogeographic analyses and analyzed mobility data from across the U.S., and found that SARS-CoV-2 in Louisiana may have originated from Texas. Prior to Mardi Gras, our analyses demonstrated that Texas is more than twice as likely as the next most probable state to be the source of SARS-CoV-2 lineages in New Orleans, while SARS-CoV-2 in Shreveport likely originated from New Orleans itself (**Figure 4A, B**).

**Figure 4.**
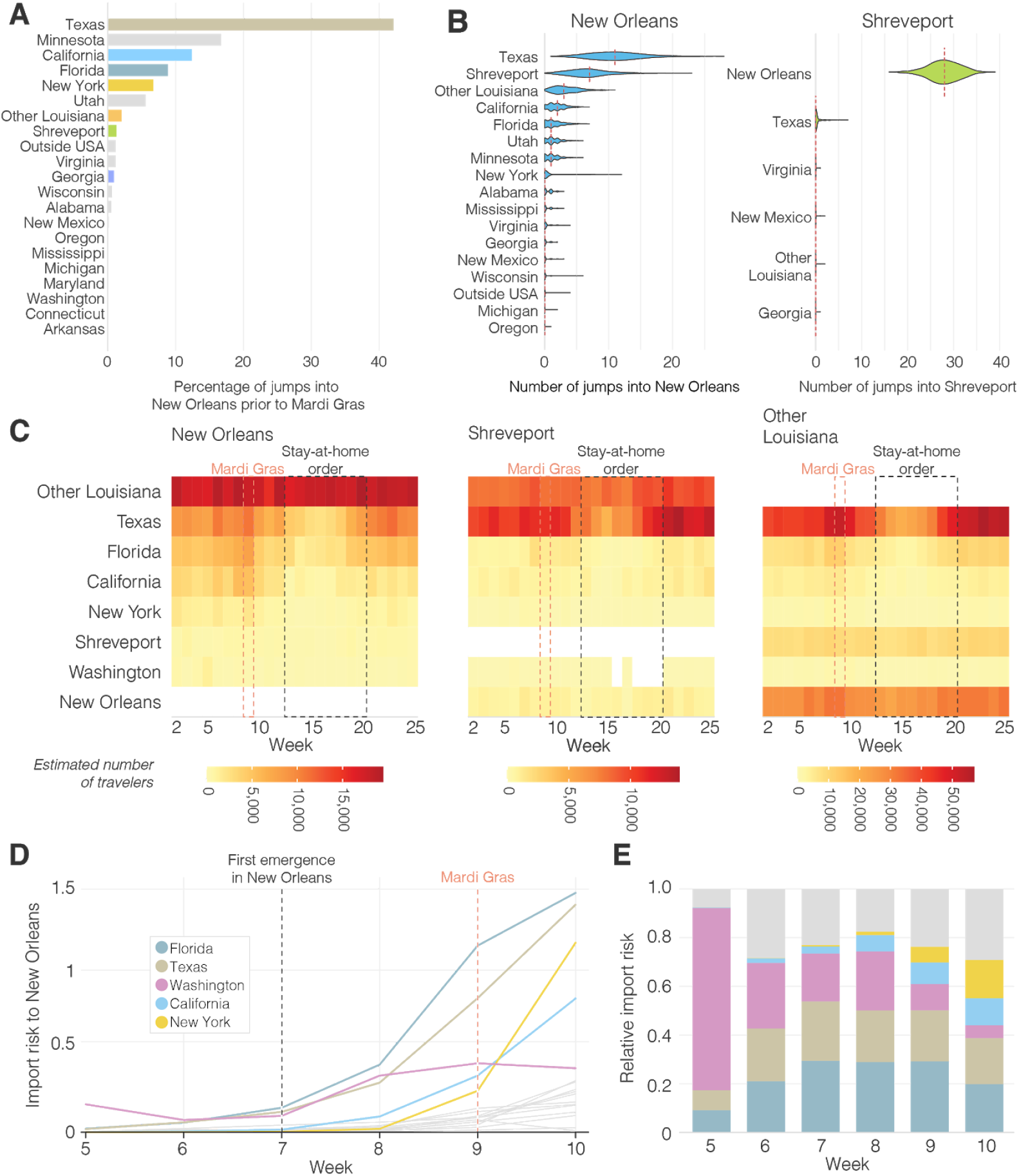
Origin of SARS-CoV-2 emergence in Louisiana. (**A**) Relative distribution of Markov jumps by origin state. Only Markov jumps that occurred before Mardi Gras day (Feb 25^th^) were included. (**B**) Estimated number of Markov jumps into New Orleans (left) and Shreveport (right). (**C**) Estimated number of travelers from states with the highest travel volumes to New Orleans, Shreveport and other parishes in Louisiana. (**D**) Import risk to New Orleans. Large Southern U.S. states and U.S. states that had early outbreaks of SARS-CoV-2 U.S. are color-coded. Other U.S. states that were included in the phylogenetic analysis are shown in grey. (**E**) Relative import risk into New Orleans. Grey area represents other U.S. states that were included in the phylogenetic analysis.

Although these analyses point to Texas as a likely source of the Louisiana clade, our phylogeographic inference is limited by geographic and temporal sampling^21^. Therefore, we also investigated movement between New Orleans, Shreveport and other U.S. states by analyzing human mobility patterns. To determine the number of travelers into Louisiana from states in the U.S. that were represented in our phylogenetic analysis, we used weekly mobility data generated by SafeGraph^22^. We found that travel movements in the week of February 13^th^ into Louisiana were strongly dominated by Texas, which accounted for 13% of travel to New Orleans, and 35% of travel to Shreveport (**Figure 4C**). These findings suggest that Texas and other regions of Louisiana were the main origins of travel into New Orleans and Shreveport during February, 2020.

To investigate the SARS-CoV-2 importation risk into New Orleans during February, 2020, we estimated the import risk based on the number of incoming travelers and the SARS-CoV-2 incidence rate at likely U.S. states of origin. We found that although the overall import risk into New Orleans was small, during the week of the likely initial introduction (February 13^th^; Week 7; **Figure 4D**), Florida and Texas represented 29% and 24% of the total import risk, respectively, whereas we estimated a lower proportion of import risk from more distant states, including California (3%), Washington (20%) and New York (0.2%; **Figure 4E**). These results are in agreement with the findings from our phylogenetic and mobility analyses, suggesting that the Louisiana clade may have originated via an introduction of SARS-CoV-2 from Texas.

### Exportation of SARS-CoV-2 from New Orleans may have caused localized outbreaks in nearby states

Our observation that superspreading during Mardi Gras likely led to increased transmission rates within New Orleans prompted us to investigate if this could also have resulted in spread to other U.S. states. We analyzed SARS-CoV-2 exports from New Orleans using mobility and genomic data in the four weeks after Mardi Gras until the stay-at-home order on March 23, which resulted in a large decline of travel and incidence. We found that the export from New Orleans was highest for nearby states and regions, in particular other parts of Louisiana, Mississippi, Alabama and Texas (**Figure 5**).

**Figure 5.**
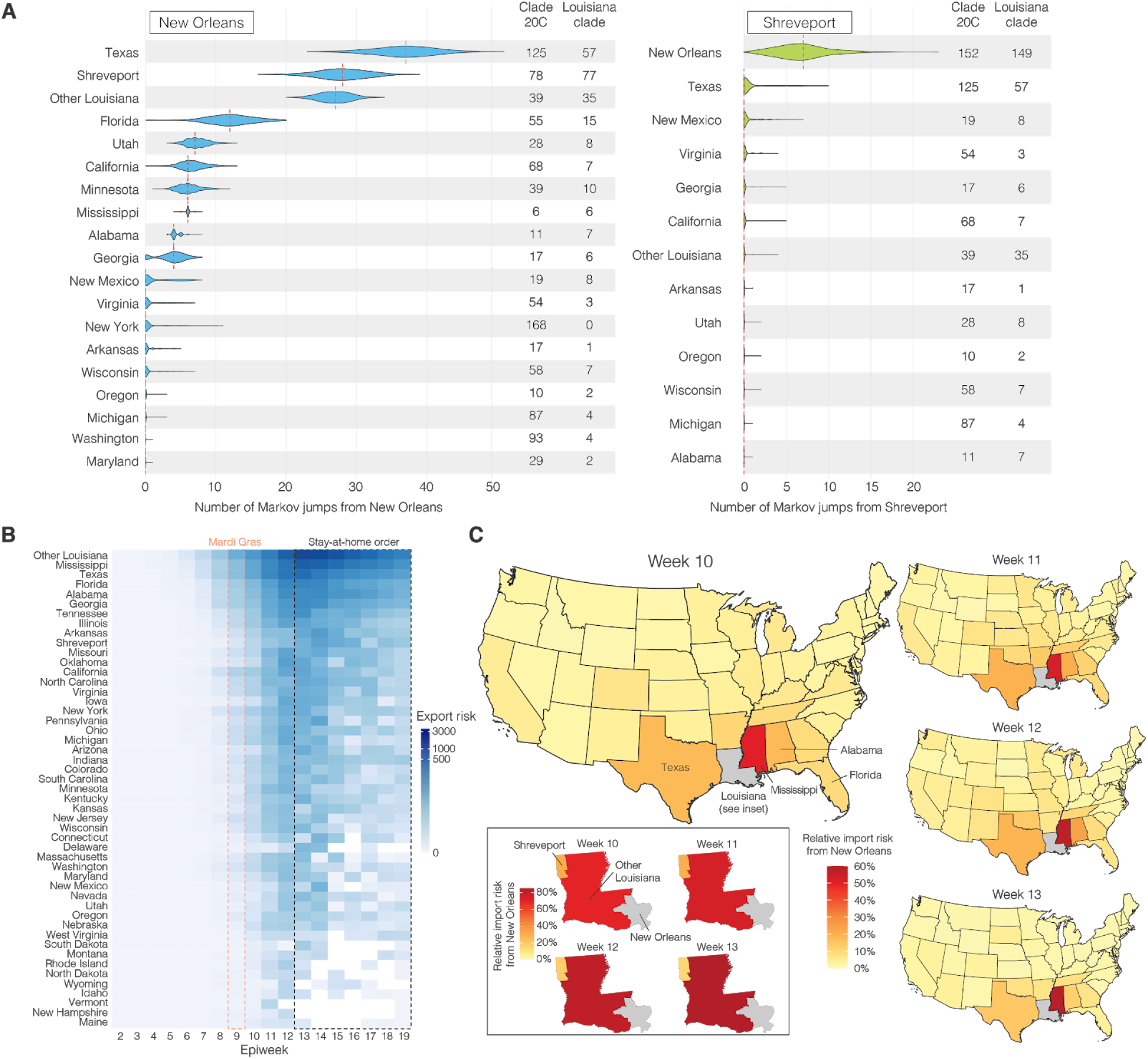
SARS-CoV-2 export risk from Louisiana. (**A**) Estimated number of Markov jumps from New Orleans (left) and Shreveport (right). On the right of each graph the number of sequences in the dataset belonging to clade 20C and the Louisiana clade are shown. (**B**) Estimated number of infected travellers from New Orleans per week. (**C**) Percentage of import risk in the lower 48 U.S. states that can be attributed to New Orleans in the four epidemiological weeks after Mardi Gras. Inset shows local relative import risk from New Orleans within Louisiana.

To determine to what extent increased transmission following superspreading during Mardi Gras could have resulted in SARS-CoV-2 infections in other states, we analyzed Markov jumps from New Orleans to regions in Louisiana and states across the U.S.. We found that SARS-CoV-2 from New Orleans may have primarily spread to nearby regions, in particular Texas and Louisiana (**Figure 5A**). In contrast, transmission in Shreveport, where we did not observe increased transmission following Mardi Gras, did not show large amounts of spread to other locations other than New Orleans (**Figure 5A**). However, since Markov Jumps from New Orleans following Mardi Gras exclusively occurred within the Louisiana clade, we compared the number of Markov jumps to the number of genomes in the Louisiana clade for each location. We found that the majority of all SARS-CoV-2 jumps into Mississippi and Alabama can be traced back to New Orleans (**Figure 5A**), suggesting that SARS-CoV-2 transmission in New Orleans may have resulted in regional spreading of COVID-19.

To further investigate to what extent increased transmission in New Orleans may have acted as a source for seeding SARS-CoV-2 to other U.S. states, we estimated the export risk from New Orleans by analyzing travel movements between New Orleans and U.S. states. We found that the export risk from New Orleans was highest to nearby regions and states, in particular to other parts of Louisiana, Mississippi and Texas (**Figure 5B**). In the four weeks between the end of Mardi Gras and the stay-at-home order, these accounted for 60% of all exported risk from New Orleans, increasing to 70% of all risk in the subsequent weeks when air travel was highly restricted (**Figure 5B**). In line with our phylogenetic analyses, we found that SARS-CoV-2 exports from Shreveport were substantially lower than from New Orleans (**Figure S2**).

As export risk from New Orleans was strongly driven by travel movements, our estimates were inherently biased towards states with larger populations. Therefore, to determine the impact of SARS-CoV-2 exports from New Orleans on local SARS-CoV-2 transmission in each U.S. state, we estimated the relative import risk from New Orleans by calculating the percentage of total SARS-CoV-2 import risk for each state that could be attributed to New Orleans. We found that the relative import risk from New Orleans was highest in neighboring U.S. states or regions (**Figure 5C**). In particular, for Mississippi and other parts of Louisiana, we found that the majority of the SARS-CoV-2 imports may have come from New Orleans (**Figure 5C**). Although the relative import risk from New Orleans declined everywhere after the statewide stay-at-home order, the decline was less pronounced for Mississippi and Louisiana, which both consistently had the highest relative import risks from New Orleans throughout the entire first wave of the COVID-19 epidemic in Louisiana (**Figures S3, S4**). Taken together, both our phylogenetic and mobility analysis suggest that the early COVID-19 epidemic in New Orleans was amplified by superspreading during Mardi Gras and may have helped seed local outbreaks in neighboring U.S. states and regions.

## Discussion

In this study, we show that domestic travel likely introduced SARS-CoV-2 into Louisiana and that a single introduction directly led to the vast majority of transmission during the first wave. Furthermore, we present several lines of evidence showing that it is likely that the Mardi Gras festival in New Orleans was a superspreading event: (i) an unusual lack of genetic diversity of SARS-CoV-2 in Louisiana, which is in sharp contrast with what has been seen in other large U.S. cities and more similar to what has been observed during cruise ship outbreaks; (ii) although our analyses suggest that SARS-CoV-2 was likely transmitting locally before Mardi Gras, we found that it is unlikely that the observed epidemiological curve in New Orleans could have been recapitulated without superspreading during Mardi Gras; and (iii) infection rates in New Orleans in the weeks immediately following Mardi Gras were substantially higher than in other major cities throughout the U.S..

The rapid nature of the early COVID-19 epidemic in New Orleans likely resulted in thousands of additional cases, which is supported by seroprevalence studies showing exposure rates of close to ten percent by May 15, 2020 in New Orleans^23^. Compared to neighboring states that did not experience the same explosive first waves as Louisiana, the CDC’s Nationwide Commercial Laboratory Seroprevalence Survey estimated that the seroprevalence in Louisiana was 35%-134% higher than in other states in the Southern U.S.^24^.

SARS-CoV-2 superspreading events can dramatically change the course of local outbreaks and have long-lasting effects. Previously, superspreading during a biotech conference in Boston in early 2020^25^ and a motorcycle rally in Sturgeon, South Dakota in August, 2020^26^ have been estimated to have resulted in more than 250,000 SARS-CoV-2 infections. Although we did not attempt to estimate the exact magnitude of the Mardi Gras superspreading event, given the lack of genetic diversity of SARS-CoV-2 within Louisiana, it seems likely that the majority of the ∼50,000 confirmed COVID-19 cases during the first wave ^27^ can be traced back to Mardi Gras.

The number of COVID-19 cases in the U.S. was severely underreported in February and March, 2020 due to a lack of adequate testing capacity. To estimate likely case numbers during this period, we modeled the number of infections based on reported COVID-19 deaths. We used a Bayesian modeling approach that was initially developed to assess the effect of non-pharmaceutical interventions in Europe^19^, and has since been widely used to reconstruct epidemiological curves in France^28^, Brazil^29^ and the UK^30^, among others. We found that reconstructed case counts by our model were slightly lower than in a serological survey in New Orleans^23^ (1.0% [our model] vs 1.6% [sero survey] of the population infected by May 15^th^), hence the scale at which superspreading occurred during Mardi Gras may have been larger than we estimated here.

We used a combination of genomic and mobility data to investigate the import and export of SARS-CoV-2 into and out of Louisiana. Our phylogenetic analyses show that SARS-CoV-2 in Louisiana most likely originated from Texas (**Figure 4**). However, most of the Louisiana clade consists of sequences from various U.S. states that either share the basal node of the Louisiana clade or belong to unresolved polytomies originating from this node. This makes accurate phylogeographic inference challenging, particularly in situations with rapid spread between different locations^31^. Previous genomic epidemiology studies investigating the emergence of SARS-CoV-2 in San Francisco^9^, Boston^32^, and New York^8^ showed that determining the source of introduction during the early stages of the COVID-19 pandemic can be challenging. A particularly illustrative example is the (re-)emergence of SARS-CoV-2 in Washington state in January/February 2020. The first case in Washington was linked to recent travel from China^33^, and when six weeks later other, genetically similar cases were detected, it was initially thought to be the result of community transmission in the context of inadequate testing^33^. Only after a reanalysis with related SARS-CoV-2 genomes from nearby British Columbia, Canada, could prolonged local transmission be excluded in favor of a more likely explanation of additional virus introduction(s) into the state^7^. In this study, we supplemented our phylogenetic analyses with large-scale analyses of travel and mobility patterns, to gain more confidence in our finding that the SARS-CoV-2 in Louisiana may have been introduced via travel from Texas. However, our estimates remain unsure and much more extensive sequencing of SARS-CoV-2 from early in the U.S. epidemic would be required to obtain more conclusive answers.

We used mobility data to determine human movement between U.S. states. Such movement, however, changed dramatically over the course of the pandemic, particularly air travel^34^. In addition, we found that air travel, as expected, can be a poor indicator of short-distance movement (**Figure S5**). To capture human movements of short distances, we therefore used weekly SafeGraph mobility data, which is based on cell phone tracking^22^. Cell phone tracking data has been shown to capture human movements on various distance scales^35,36^. To further increase the accuracy of our mobility analysis and mitigate large swings in human movements due to government intervention, we only analyzed travel until mid-March, before Louisiana and many other states adopted stay-at-home orders, and travel substantially decreased.

Our phylogenetic analyses indicate that SARS-CoV-2 was introduced into New Orleans multiple times, but that only one main clade (the “Louisiana clade”) was eventually successful in establishing widespread community transmission. We estimated that the emergence of the Louisiana clade in New Orleans occurred in mid-February, just prior to Mardi Gras. However, estimating an accurate introduction date with limited genetic diversity can be challenging^16^. We therefore investigated timing by estimating both the time of introduction by analyzing Markov jumps and the start of local transmission by determining the TMRCA of the Louisiana clade. We found that both analyses suggest that the Louisiana clade was likely present in New Orleans prior to Mardi Gras.

With the recent emergence of more transmissible SARS-CoV-2 variants in the U.S.^37^ and elsewhere^38^, robust virus genomic surveillance systems and analysis frameworks will be critical to provide insights into the ongoing spread and evolution of SARS-CoV-2. We show that a single introduction of SARS-CoV-2 can rapidly find its way through an unprotected population and cause large-scale epidemics in the absence of adequate testing and control efforts. Our study provides a key example of how a large-scale event played a key role during the early epidemic in the U.S. and how such events may continue to play a role in amplifying local outbreaks if SARS-CoV-2 is left unchecked.

## Methods

### Ethics Statement

Sample collection, RNA extraction, and viral sequencing was evaluated by the Institutional Review Boards (IRBs) at Tulane University (IRB# 2020-396), Louisiana State University Health System (LSUHS) (IRB# STUDY00001445) and Ochsner Health (IRB# 2019.334). All samples were de-identified before receipt by the study investigators.

### Sample Collection and RNA extraction

Nasopharyngeal swabs from Tulane Medical Center were collected March-April 2020 from 1) hospitalized COVID-19 patients consenting to participate in viral isolation and sequencing studies and 2) left-over clinical samples from individuals presenting to the Emergency Department (ED) with COVID-19 symptoms. Nasopharyngeal swabs from LSUHS and Ochsner health were left-over clinical samples from either outpatient or hospitalized individuals.

Viral RNA was extracted using the QIAmp Viral RNA Mini Kit (Qiagen), Quick-RNA Viral Kit (Zymo Research) or Mag-Bind Viral DNA/RNA kit (Omega Bio-tek) according to the manufacturer’s instructions. RNA extracts from samples collected at Tulane Medical Center were screened for presence of SARS-CoV-2 Nucleocapsid gene according to the 2019-nCoV Real Time rRT-PCR Panel protocol^39^ on the QuantStudio 3 (Applied Biosciences); only the N1 Primer/Probe Mix was used(F: 5’-GACCCCAAAATCAGCGAAAT-3’, R: 5’-TCTGGTTACTGCCAGTTGAATCTG-3’, Probe: 5’-FAM-ACCCCGCAT/ZEN/TACGTTTGGTGGACC-3IABkFQ-3’). Samples with a Ct<30 (correlating to ∼500 copies of virus/µL) were selected for amplicon sequencing and viral RNA was shipped to Scripps Research Institute. RNA extracts from samples collected at LSUHS were screened with an EUA diagnostic RT-qPCR at the LSUHS emerging viral threat laboratory and shipped for sequencing to the Microbial Genome Sequencing Center (MiGS) in Pittsburgh, PA.

### SARS-CoV-2 Amplicon Sequencing

SARS-CoV-2 was sequenced using PrimalSeq-Nextera XT. This protocol is based on the ARTIC PrimalSeq protocol and adapted for Illumina Nextera XT library preparation^40^. The ARTIC network nCoV-2019 V3 primer scheme uses two multiplexed primer pools to create overlapping 400 bp amplicon fragments in two PCR reactions. Instead of ligating Illumina adapters, Nextera XT is used to circumvent the 2×250 or 2×300 read length requirement. A detailed version of this protocol can be found here: https://andersen-lab.com/secrets/protocols/. Briefly, SARS-CoV-2 RNA (2 µL) was reverse transcribed with SuperScript IV VILO. The virus cDNA was amplified in two multiplexed PCR reactions (one reaction per ARTIC network primer pool) using Q5 DNA High-fidelity Polymerase (New England Biolabs). Following an AMPureXP bead (Beckman Coulter) purification of the combined PCR products, the amplicons were diluted and libraries were prepared using Nextera XT (Illumina) or NEBNext Ultra II DNA Library Prep Kits (New England Biolabs). The libraries were purified with AMPureXP beads and quantified using the Qubit High Sensitivity DNA assay kit (Invitrogen) and Tapestation D5000 tape (Agilent). The individual libraries were normalized and pooled in equimolar amounts at 2 nM. The 2 nM library pool was sequenced on an Illumina NextSeq using a 500/550 Mid Output Kit v2.5 (300 Cycles). A subset of samples from Ochsner Health were processed without tagmentation and sequenced on a Illumina MiSeq using a MiSeq reagent kit V3 (600 cycles). Raw reads were deposited under BioProject accession ID’s PRJNA643575 and PRJNA612578.

Consensus sequences were assembled using an inhouse Snakemake ^41^ pipeline with bwa-mem ^42^ and iVar v1.2.2 ^42,43^.

### SARS-CoV-2 metagenomic Sequencing

For samples that were collected at Ochsner Health we used the following metagenomic sequencing protocol: RNA isolated from VTM was converted to double stranded cDNA and sequencing libraries prepared using TruSeq Stranded RNA Library Preparation Kit (Illumina) according to the manufacturer’s instructions. The sequencing libraries were evaluated using high sensitivity D5000 ScreenTape in the 4200 TapeStation system (Agilent) and quantified using Library Quantitation Kit (Roche). The libraries normalized and pooled, and subsequently sequenced using the NextSeq and 500/550 2×150 MID Output format (Illumina). Raw reads were deposited under BioProject accession ID PRJNA643574.

For samples that were collected at LSUHS we used the following metagenomic sequencing protocol: For each sample, 13µL of extracted RNA was reverse transcribed using the Maxima H-minus ds cDNA kits (ThermoFisher Scientific). Libraries were enriched using a Nextera Flex for Enrichment Library Preparation kit with a Respiratory Virus Oligo Set v1, with samples being pooled in 12-plex enrichment reactions. The resulting pools were quantified and grouped in sets of no more than 48 samples and run on a NextSeq 550 using a 150cyc High Output Flow Cell. We used BreSeq^44^ (v.0.34.1) to map reads to Wuhan-Hu-1 SARS-CoV-2 (NC_045512) or 2019-nCoV WIV04 (EPI_ISL_402124)^2^ and call the consensus sequence. All predicted mutations were reported for isolates exceeding mean 40x coverage. Raw reads were deposited under BioProject accession ID PRJNA681020.

### Phylogenetic Analysis

We used the global SARS-CoV-2 phylogeny provided by Rob Lanfear^45^ as of Oct 21^st^ from GISAID (**Table S2**) and narrowed it down to 1,171 full-length genomes representing the genetic diversity from 19 different states in the USA and 228 sequences from outside the USA. The number of genomes from each state are shown in **Table S3**. We also masked sites in the alignment that were homoplastic as shown in **Table S4**. We used this dataset to estimate a starting tree using a HKY^46^ nucleotide substitution model, with a strict clock model using a non-informative continuous-time Markov chain (CTMC) reference prior ^47^ and an exponential population prior implemented in BEAST v1.10.5pre^17^. We used the maximum clade credibility tree from this analysis as a starting tree to estimate the movement of the virus between geographic locations under a flexible discrete-state phylogeographic framework^48^ using BEAST v1.10.5pre^17^. We used a HKY nucleotide substitution model under an uncorrelated relaxed clock model^49^, an exponential population prior and a symmetric discrete-state substitution model. We included a Markov jump counting procedure^18^ to estimate the number of specific transitions between locations while simultaneously accounting for the large uncertainty in phylogenetic reconstruction. Specifically, to characterize the proportion of introductions from each discrete state into New Orleans and Shreveport, we first compute the relative number of the earliest Markov jump from each discrete state to New Orleans or Shreveport along the phylogenetic tree for each posterior sample. We then summarize these proportions over all samples to learn their posterior distributions. We simulated two independent MCMC chains for 100 million steps each and discarded the first 10 million steps as burnin in each. Effective sample sizes for scientifically relevant model parameters were all above 200. The BEAST XML and log files are available at https://github.com/andersen-lab/paper_2020_new-orleans-hcov-genomics.

### Travel data

We calculated travel between counties using the weekly patterns data from SafeGraph^50^ a data company that aggregates anonymized location data from numerous applications in order to provide insights about physical places, via the Placekey^51^ Community. To enhance privacy, SafeGraph excludes census block group information if fewer than five devices visited an establishment in a month from a given census block group. We estimated the true number of travellers for a given week, *w*, between a source census block group, *cbgs* and a destination census block group, *cbgd* (V_w,cbgs,cbgd_) using the raw number of visitor counts for week, *w*, identified from points of interest in *cbgd* from *cbgs* (C_w,cbgs,cbgd_), the total number of visitors with a known source census block group and the population of *cbgd*, according to 

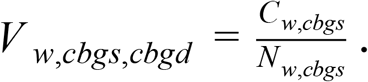

We also obtained monthly air travel passenger data between the 19 U.S. states from the International Air Transportation Association. We used Apache Spark v2.4.6 and PySpark v2.4.6 to preprocess data from SafeGraph to estimate the travel between states.

There was a strong correlation in travel trends between mobility data and air travel passenger counts, but unlike SafeGraph mobility data, air travel data was unable to capture travel over short distances (R^2^=0.80; **Figure S5**). The code used to estimate movement between states using mobility data is available at https://github.com/andersen-lab/paper_2020_new-orleans-hcov-genomics.

### Incidence

We used the R package Epidemia^19^ to estimate the number of infections over time for each state and metro area independently using the number of deaths. Epidemia estimates a time-varying reproduction number, *R*_*t*_ from the observed number of deaths, informed by an infection-to-death distribution and infection fatality rate estimate. We obtained the number of deaths and regions through the outbreak.info R package^52^, which aggregates epidemiological data from the COVID-19 data repository by the Center for Systems Science and Engineering (CSSE) at Johns Hopkins University ^53^ and the COVID-19 data repository by the New York Times (https://github.com/nytimes/covid-19-data). The code used to estimate the number of infections is available at https://github.com/andersen-lab/paper_2020_new-orleans-hcov-genomics.

With the modeled infection curve we accurately reconstructed the daily number of COVID-19 deaths as well as the infection fatality rate (1.0%; 95% HPD: [0.7%, 1.3%] versus 1.6% on May 15^th^ according to ^23^). We calculated the number of expected cases based on 100,000 simulations of a negative binomial branching process model. Following Lloyd-Smith *et al*.^20^, we assumed that secondary infections from a single infection would follow a negative binomial distribution described by an *R*_0_ of 2.6 and the overdispersion parameter, *k* of 0.16. In addition, we assumed that the local transmission in New Orleans started with a single introduction of the virus on February 13^th^ (median TMRCA of Louisiana clade) and a serial interval of 4 days. The code to run the branching process model is available at https://github.com/andersen-lab/paper_2020_new-orleans-hcov-genomics.

### Import/export risk

We assumed a gamma distributed incubation period with shape 5.807 and rate 1.055^54^. We estimated the number of cases that started showing symptoms using 

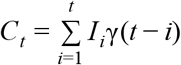

where γ(*t* − *i*) is the probability distribution function of the incubation period and *I*_*i*_ is the estimated number of infections on a given day, *i*.

We assumed that cases were infectious one day before symptom onset and a gamma distributed infectious period with shape 2.5 and rate 0.35^55^. As per Fauver *et al*.^*56*^, we assumed that cases would not travel after receiving a positive clinical test and we retrieved the number of confirmed cases as reported by state and local health departments through the outbreak.info R package to obtain the reported number of cases. We assumed a uniform ascertainment period of 5 days for the reported cases. We estimated the number of infectious cases that could travel on a given day, *t*, using 

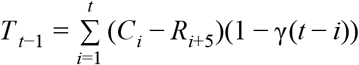

where γ(*t* − *i*) is the cumulative distribution function of the infectious period and *C*_*i*_ is the number of cases that start showing symptoms on a given day, *i*, and *R*_*i*_ is the number of reported cases on day, *i*.

We estimated the number of infectious travellers coming into a destination, *d*, from a source, *s*, using 

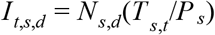

where *P* _*s*_ is the population at the source, *T* _*s,t*_ is the number of infectious cases that could travel at the source and *N*_*s,d*_ is the number of travellers from the source to the destination. We used this estimate to compare importation and exportation risk. The code to estimate the import and export risk is available at https://github.com/andersen-lab/paper_2020_new-orleans-hcov-genomics.

## Data Availability

The data that support the findings of this study are openly available in Github at https://github.com/andersen-lab/paper_2020_new-orleans-hcov-genomics

## Acknowledgements

We thank the administrators of the GISAID database for supporting rapid and transparent sharing of genomic data during the COVID-19 pandemic and to all our colleagues sharing data on GISAID. A full list acknowledging the authors submitting genome sequence data used in this study can be found in **Table S2**. The research leading to these results has received funding from the National Institutes of Health (grants U19AI135995, 3U19AI135995-03S2, UL1TR002550, U01AI151812, U01AI124302, R01AI153044 and R01HG006139), the European Research Council under the European Union’s Horizon 2020 research and innovation program (grant agreement no. 725422-ReservoirDOCS) and from the European Union’s Horizon 2020 project MOOD (grant agreement no. 874850). The Artic Network receives funding from the Wellcome Trust through project 206298/Z/17/Z. P.L. acknowledges support by the Research Foundation—Flanders (“Fonds voor Wetenschappelijk Onderzoek—Vlaanderen”, G066215N, G0D5117N, and G0B9317N). S.L.L. acknowledges support by the National Science Foundation Small Business Innovation Research grants 2027424 and 1830867. We also gratefully acknowledge support from NVIDIA Corporation and Advanced Micro Devices, Inc., with the donation of parallel computing resources used for this research.

## Contributions

M.Z., C.A., A.R.S., D.J.N, G.S-S., A.R.B-K., P.S., L.I.M., K.J.G., K.C., D.J.S., R.R.S., R.K. guided and/or performed lab experiments and prepared samples from sequencing. M.Z., K.G., D.J.S, M.A., M.M., R.R., V.S.C., P.L., M.A.S. performed data management, processing and analysis. C.A., M.McG., S.T., E.S., L.N. provided project management. A.R.S., J.A.V., R.S.S., J.G-D., A.K.F., A.D., D.F. guided research activities at clinical sites. G.D., A.W., N.L.M., L.D.H., M.A., P.B-F., P.D., S.S., C.M., N.G., S.A., G.Y., L.F., C.A.H, L.L, R.K., E.H, K.K. provided critical insights and/or study guidance. M.Z., K.G., C.A., P.L., J.P.K., S.L.L., L.G., R.F.G., M.A.S., K.G.A. oversaw study design, implementation, analysis, and drafted and revised the manuscript. All authors contributed to interpreting and reviewing the manuscript

## Supplemental Data

**Figure S1.**
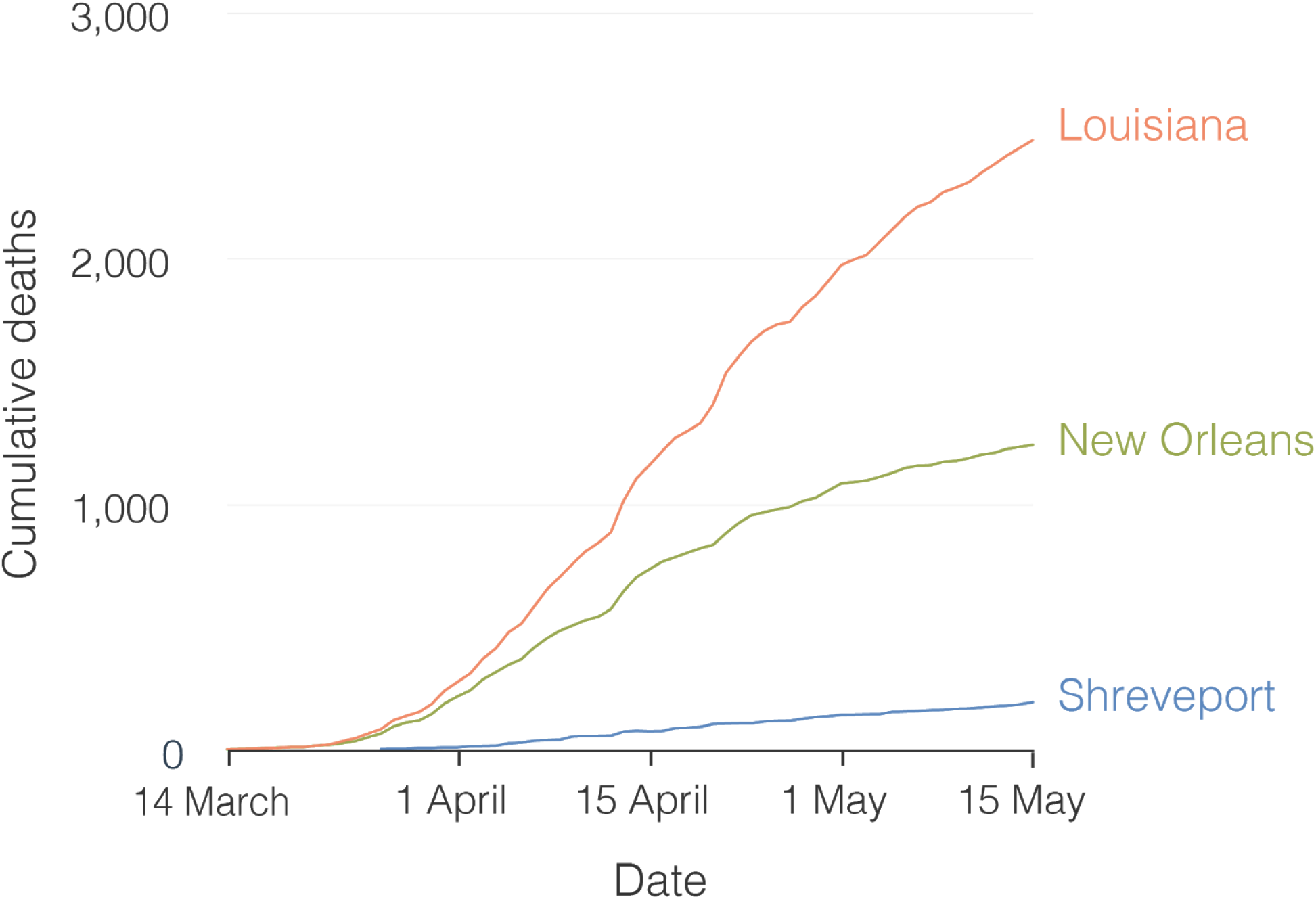
Cumulative SARS-CoV-2 death during the first wave of the COVID-19 epidemic in Louisiana. https://bit.ly/2MF5u5x

**Figure S2.**
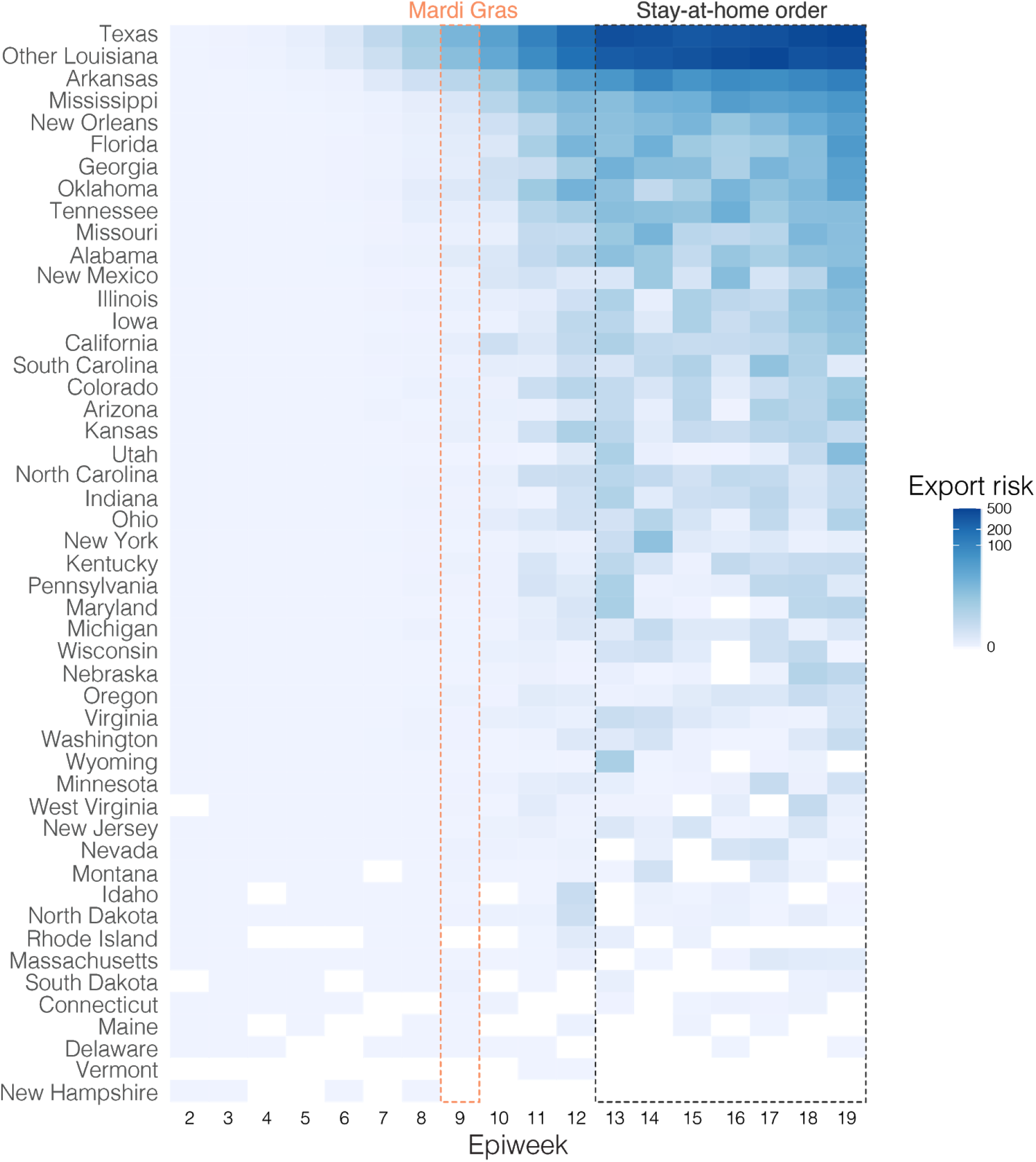
Estimated number of travelers from Shreveport per epiweek. https://bit.ly/2YMZIRN

[Animation]

**Figure S3.** Relative import risk from New Orleans for each U.S. state per epiweek.

https://bit.ly/36MTciv

[Animation]

**Figure S4.** Relative import risk from New Orleans for Shreveport and other parts of Louisiana per epiweek.

https://bit.ly/3cMDvfc

**Figure S5.**
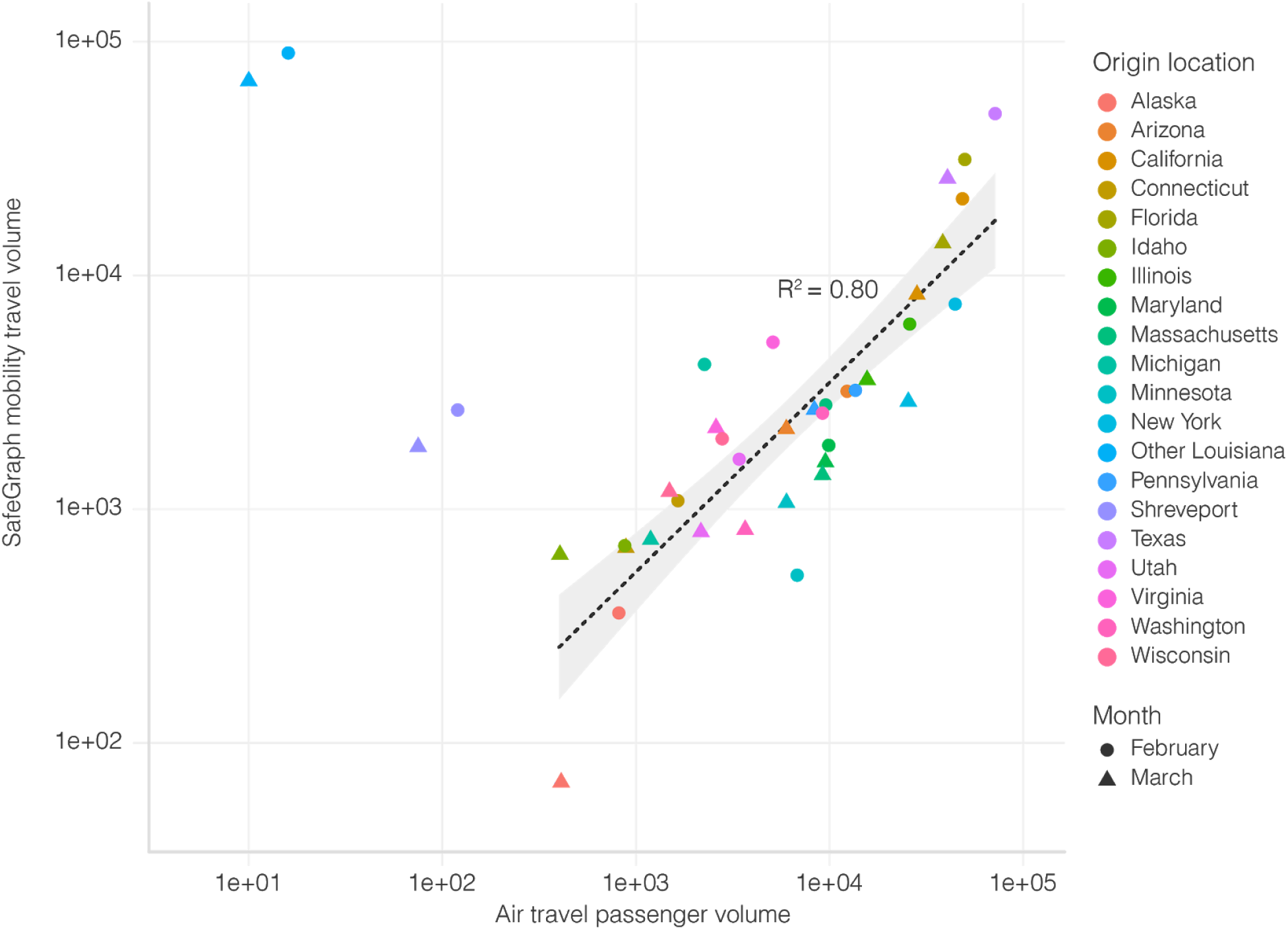
Correlation between travel datasets. Air travel passenger volumes and SafeGraph mobility travel volumes from various U.S. states into New Orleans. Spearman rank correlation does not include Shreveport and Other Louisiana, since air travel is not the dominant mode of transport to New Orleans for these locations. https://bit.ly/2YOA7If

**Table S1**. Newspaper articles about the role of Mardi Gras in the spread of SARS-CoV-2 in Louisiana.

https://bit.ly/2MWXvAS

**Table S2**. GISAID acknowledgement table.

https://bit.ly/3aBAuLO

**Table S3**. Number of sequences from each location in the genomic dataset.

https://bit.ly/39SgG7E

**Table S4**. Masked homoplasic mutations in the alignment.

https://bit.ly/3oTvl70

## References

1. Wu, F. et al. A new coronavirus associated with human respiratory disease in China. Nature vol. 579 265–269 (2020).

2. Zhou, P. et al. A pneumonia outbreak associated with a new coronavirus of probable bat origin. Nature 579, 270–273 (2020).

3. [No title]. https://www.who.int/docs/default-source/coronaviruse/situation-reports/20200123-sitrep-3-2019-ncov.pdf?sfvrsn=d6d23643_8.

4. [No title]. https://www.who.int/docs/default-source/coronaviruse/situation-reports/20200229-sitrep-40-covid-19.pdf?sfvrsn=849d0665_2.

5. First Travel-related Case of 2019 Novel Coronavirus Detected in United States. https://www.cdc.gov/media/releases/2020/p0121-novel-coronavirus-travel-case.html (2020).

6. CDC Confirms Person-to-Person Spread of New Coronavirus in the United States. https://www.cdc.gov/media/releases/2020/p0130-coronavirus-spread.html (2020).

7. Worobey, M. et al. The emergence of SARS-CoV-2 in Europe and North America. Science 370, 564–570 (2020).

8. Maurano, M. T. et al. Sequencing identifies multiple early introductions of SARS-CoV-2 to the New York City region. Genome Res. (2020) doi:10.1101/gr.266676.120.

9. Deng, X. et al. Genomic surveillance reveals multiple introductions of SARS-CoV-2 into Northern California. Science 369, 582–587 (2020).

10. Davis, J. T. et al. Estimating the establishment of local transmission and the cryptic phase of the COVID-19 pandemic in the USA. medRxiv 2020.07.06.20140285 (2020).

11. Perkins, A. et al. Estimating unobserved SARS-CoV-2 infections in the United States. medRxiv 2020.03.15.20036582 (2020).

12. Lu, F. S., Nguyen, A. T., Link, N. B., Lipsitch, M. & Santillana, M. Estimating the Early Outbreak Cumulative Incidence of COVID-19 in the United States: Three Complementary Approaches. medRxiv (2020) doi:10.1101/2020.04.18.20070821.

13. Team, C. C.-19 R. et al. Evidence for Limited Early Spread of COVID-19 Within the United States, January–February 2020. MMWR. Morbidity and Mortality Weekly Report vol. 69 680–684 (2020).

14. outbreak.info. https://outbreak.info/.

15. Sekizuka, T. et al. Haplotype networks of SARS-CoV-2 infections in the cruise ship outbreak. Proc. Natl. Acad. Sci. U. S. A. 117, 20198–20201 (2020).

16. Grubaugh, N. D. et al. Tracking virus outbreaks in the twenty-first century. Nat Microbiol 4, 10–19 (2019).

17. Suchard, M. A. et al. Bayesian phylogenetic and phylodynamic data integration using BEAST 1.10. Virus Evol 4, vey016 (2018).

18. Minin, V. N. & Suchard, M. A. Counting labeled transitions in continuous-time Markov models of evolution. J. Math. Biol. 56, 391–412 (2008).

19. Flaxman, S. et al. Estimating the effects of non-pharmaceutical interventions on COVID-19 in Europe. Nature 584, 257–261 (2020).

20. Lloyd-Smith, J. O., Schreiber, S. J., Kopp, P. E. & Getz, W. M. Superspreading and the effect of individual variation on disease emergence. Nature 438, 355–359 (2005).

21. Bloomquist, E. W., Lemey, P. & Suchard, M. A. Three roads diverged? Routes to phylogeographic inference. Trends Ecol. Evol. 25, 626–632 (2010).

22. Weekly Patterns (v2). https://docs.safegraph.com/docs/weekly-patterns.

23. Feehan, A. K. et al. Seroprevalence of SARS-CoV-2 and Infection Fatality Ratio, Orleans and Jefferson Parishes, Louisiana, USA, May 2020. Emerg. Infect. Dis. 26, 2766–2769 (2020).

24. CDC. COVID-19 Cases, Deaths, and Trends in the US. https://covid.cdc.gov/covid-data-tracker (2020).

25. Lemieux, J. E. et al. Phylogenetic analysis of SARS-CoV-2 in Boston highlights the impact of superspreading events. Science (2020) doi:10.1126/science.abe3261.

26. [No title]. http://ftp.iza.org/dp13670.pdf.

27. Outbreak.info - Louisiana. https://outbreak.info/epidemiology?location=USA_US-LA&log=false&variable=confirmed&xVariable=date&fixedY=false&percapita=false.

28. Pullano, G. et al. Underdetection of cases of COVID-19 in France threatens epidemic control. Nature 590, 134–139 (2020).

29. Candido, D. S. et al. Evolution and epidemic spread of SARS-CoV-2 in Brazil. Science 369, 1255–1260 (2020).

30. Volz, E. et al. Evaluating the Effects of SARS-CoV-2 Spike Mutation D614G on Transmissibility and Pathogenicity. Cell 184, 64–75.e11 (2021).

31. Villabona-Arenas, C. J., Hanage, W. P. & Tully, D. C. Phylogenetic interpretation during outbreaks requires caution. Nature Microbiology 5, 876–877 (2020).

32. Lemieux, J. et al. Phylogenetic analysis of SARS-CoV-2 in the Boston area highlights the role of recurrent importation and superspreading events. medRxiv (2020) doi:10.1101/2020.08.23.20178236.

33. Bedford, T. et al. Cryptic transmission of SARS-CoV-2 in Washington state. Science 370, 571–575 (2020).

34. TSA checkpoint travel numbers (current year(s) versus prior year/same weekday). https://www.tsa.gov/coronavirus/passenger-throughput.

35. Kraemer, M. U. G. et al. The effect of human mobility and control measures on the COVID-19 epidemic in China. Science 368, 493–497 (2020).

36. Chang, S. et al. Mobility network models of COVID-19 explain inequities and inform reopening. Nature 589, 82–87 (2020).

37. Galloway, S. E. Emergence of SARS-CoV-2 B.1.1.7 Lineage — United States, December 29, 2020–January 12, 2021. MMWR Morb. Mortal. Wkly. Rep. 70, (2021).

38. Volz, E. et al. Transmission of SARS-CoV-2 Lineage B.1.1.7 in England: Insights from linking epidemiological and genetic data. medRxiv 2020.12.30.20249034 (2021).

39. [No title]. https://www.fda.gov/media/134922/download.

40. Quick, J. et al. Multiplex PCR method for MinION and Illumina sequencing of Zika and other virus genomes directly from clinical samples. Nat. Protoc. 12, 1261–1276 (2017).

41. Köster, J. & Rahmann, S. Snakemake—a scalable bioinformatics workflow engine. Bioinformatics 28, 2520–2522 (2012).

42. Li, H. Aligning sequence reads, clone sequences and assembly contigs with BWA-MEM. (2013).

43. Grubaugh, N. D. et al. An amplicon-based sequencing framework for accurately measuring intrahost virus diversity using PrimalSeq and iVar. Genome Biol. 20, 1–19 (2019).

44. Deatherage, D. E. & Barrick, J. E. Identification of mutations in laboratory-evolved microbes from next-generation sequencing data using breseq. Methods Mol. Biol. 1151, 165–188 (2014).

45. roblanf & Mansfield, R. roblanf/sarscov2phylo: 13-11-20. (2020) doi:10.5281/zenodo.4289383.

46. Hasegawa, M., Kishino, H. & Yano, T. Dating of the human-ape splitting by a molecular clock of mitochondrial DNA. J. Mol. Evol. 22, 160–174 (1985).

47. Ferreira, M. A. R. & Suchard, M. A. Bayesian analysis of elapsed times in continuous-time Markov chains. Canadian Journal of Statistics vol. 36 355–368 (2008).

48. Lemey, P., Rambaut, A., Drummond, A. J. & Suchard, M. A. Bayesian phylogeography finds its roots. PLoS Comput. Biol. 5, e1000520 (2009).

49. Drummond, A. J., Ho, S. Y. W., Phillips, M. J. & Rambaut, A. Relaxed Phylogenetics and Dating with Confidence. PLoS Biol. 4, e88 (2006).

50. Places Data & Foot-Traffic Insights. https://www.safegraph.com/.

51. Placekey. https://placekey.io.

52. outbreak-info. outbreak-info/R-outbreak-info. https://github.com/outbreak-info/R-outbreak-info.

53. Dong, E., Du, H. & Gardner, L. An interactive web-based dashboard to track COVID-19 in real time. Lancet Infect. Dis. (2020) doi:10.1016/S1473-3099(20)30120-1.

54. Lauer, S. A. et al. The Incubation Period of Coronavirus Disease 2019 (COVID-19) From Publicly Reported Confirmed Cases: Estimation and Application. Ann. Intern. Med. 172, 577–582 (2020).

55. Jung, S.-M. et al. Real-Time Estimation of the Risk of Death from Novel Coronavirus (COVID-19) Infection: Inference Using Exported Cases. Epidemiology 330 (2020).

56. Fauver, J. R. et al. Coast-to-Coast Spread of SARS-CoV-2 during the Early Epidemic in the United States. Cell 181, 990–996.e5 (2020).

