## Supplementary figures and images for "Emergence of an early SARS-CoV-2 epidemic in the United States"

### Figure S1

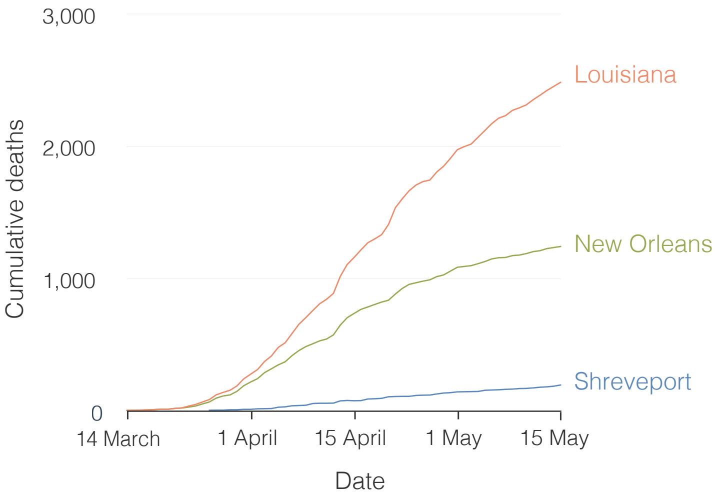

### Figure S2

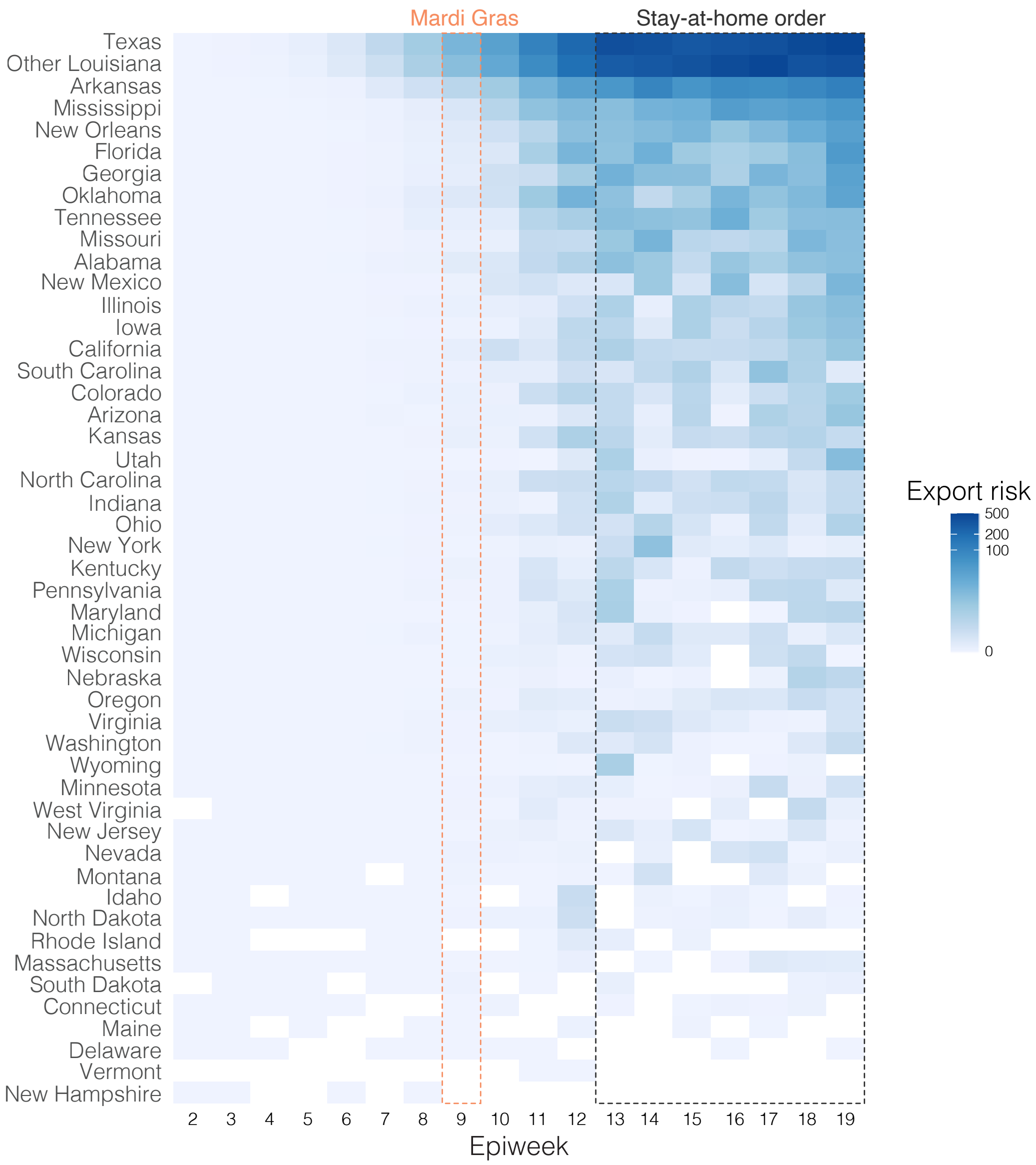

### Figure S5

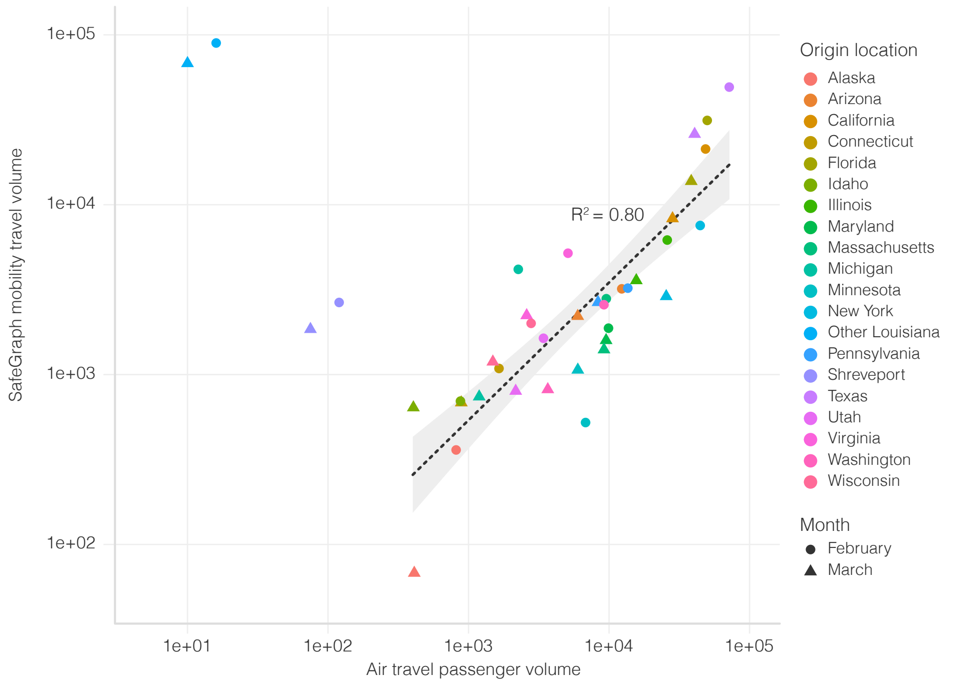
